# Multi-modal and multi-organ *in vivo* imaging to assess geroprotective interventions in humans: results from a pilot trial of rapamycin in Alzheimer’s Disease

**DOI:** 10.64898/2026.03.11.26348179

**Authors:** Pontus Plavén-Sigray, Martin Bolin, Emilia Palmér, Ruben Dörfel, Daniel Thor, Martin Schain, Maria Nilsson, Navid Golpour, Rune Brautaset, Pete A Williams, Rubens Spin-Neto, Monika Probst, Jenny Castaings, Peder Sörensson, David Marlevi, Marcus Carlsson, Miia Kivipelto, Jonas E. Svensson

## Abstract

**Background:** Geroprotective interventions, including the mTOR inhibitor rapamycin, slow aging in preclinical models. Translation to humans remains challenging because clinical trials require endpoints detectable within feasible timeframes. Multi-modal *in vivo* imaging could address this limitation by enabling simultaneous assessment of age-related pathology across multiple organ systems, but its feasibility in clinical trials is uncertain.

**Objective:** To evaluate the feasibility of deploying a multi-modal, multi-organ imaging battery in a geroprotective intervention trial of rapamycin and to collect exploratory efficacy data across multiple domains of age-related pathology.

**Methods:** In a single-center, open-label, single-arm pilot trial, 14 participants with early-stage Alzheimer’s disease (MCI or mild dementia; Montreal Cognitive Assessment ≥18; amyloid-positive) received oral rapamycin 7 mg once weekly for 26 weeks. Participants underwent baseline and end-of-treatment imaging including retinal optical coherence tomography (OCT); [^18^F]FDG positron emission tomography/computed tomography (PET/CT) of the head, thorax, and lower spine; dentomaxillofacial MRI; and cardiac MRI with stress perfusion and arterial pulse wave velocity. Feasibility outcomes included completion rates and technical or logistical barriers. Exploratory pre-post changes were assessed using paired t-tests.

**Results:** Of the 14 enrolled participants, 13 completed follow-up imaging. Among these, completion was 100% for OCT, [^18^F]FDG PET/CT, and dentomaxillofacial MRI. Cardiac MRI and pulse wave velocity were completed in 69% (9/13), primarily limited by scanner access during a healthcare worker strike. No imaging-related adverse events occurred. Exploratory analyses showed nominally significant pre-post increases in cardiac output (p=0.017), late diastolic (A-wave) kinetic energy (average: p=0.044; peak: p=0.024), left retinal ganglion cell layer thickness (p=0.044), and optic nerve head [^18^F]FDG uptake (p=0.040). Bone mineral density showed no significant pre-post changes, while muscle cross-sectional area decreased numerically but not significantly (p=0.058). In exposure-response analyses, higher rapamycin blood concentration was significantly correlated with greater skeletal muscle density (r=0.64, p=0.035) and, albeit not significantly, smaller loss of cross-sectional area (r=−0.53, p=0.097).

**Conclusions:** A multi-modal imaging battery spanning several organ systems was successfully integrated into a clinical trial, with high completion rates for most modalities. Logistical constraints were the primary barriers affecting cardiac measures. These findings inform the design of future randomized trials of geroprotective interventions, where such imaging batteries may help detect changes in age-related pathology over relatively short timeframes.

## 1. Introduction

A growing number of geroprotective interventions (GPIs) have demonstrated promising effects on lifespan and healthspan by delaying the onset of age-related mortality and morbidity in preclinical models (1,2). However, few interventions have been systematically evaluated in humans (3). A major barrier to translation is long human lifespan, which makes mortality or disease incidence impractical as primary endpoints in early-phase trials. This has prompted the search for surrogate endpoints that can detect geroprotective effects within feasible trial durations (4,5). Because most candidate GPIs are expected to act on fundamental aging mechanisms, their beneficial effects should manifest across multiple tissues and physiological systems affected by age. An effective GPI will inhibit or slow age-related pathological processes throughout the body, rather than targeting a single disease pathway (4,6). This distinguishes GPIs from conventional disease-modifying interventions and suggests that efficacy assessments must capture changes in aging pathology across different tissue types and systems (7,8).

We recently proposed the use of *in vivo* multimodal medical imaging to evaluate candidate GPIs by detecting changes across a broad range of age-related pathological processes before the clinical onset of disease (9). Imaging enables visualization and objective quantification of key age-related pathologies, such as atherosclerosis, loss of bone mineral density and skeletal muscle mass, retinal neurodegeneration, and cardiac remodelling. These processes are common in aging populations, increase with age, and are often detectable years before overt clinical disease. A battery of tests that integrates *in vivo* imaging protocols for these processes could therefore provide a diverse set of outcome measures reflecting the pace of general age-related pathology accumulation across the body, enabling efficient screening of candidate GPIs within manageable time frames in early-phase trials. However, the practical feasibility of deploying such a multimodal, multi-organ imaging battery in a GPI clinical trial setting has not yet been demonstrated.

Rapamycin (*sirolimus*), an inhibitor of the mechanistic target of rapamycin (mTOR), represents a promising candidate for testing this imaging-based approach. The compound has the strongest and most replicated preclinical evidence for geroprotection in multiple independent studies, demonstrating lifespan extension in a range of model organisms (10), including mammals (11). Rapamycin has also demonstrated beneficial effects in preclinical models across different organ systems that are assessable by medical imaging, such as increased cardiac function (12), increased retinal neuroprotection (13), decreased periodontal disease (14,15), reduced age-related bone loss (16), and reversal of neurodegenerative pathology (17); with strong preclinical evidence for a beneficial effect in Alzheimer’s disease (AD) (18).

To assess the feasibility of deploying a multi-modal, multi-organ *in vivo* imaging battery for assessing candidate GPIs in humans, we conducted a clinical pilot trial (19): Participants with early-stage AD received oral rapamycin (7 mg weekly) for six months, with multimodal imaging of both the central nervous system and peripheral organ systems performed at baseline and at the end of treatment.

The selection of an early-stage AD population served dual purposes. First, rapamycin has demonstrated robust beneficial effects in preclinical models of neurodegeneration (17) with AD repeatably suggested as a promising indication for rapamycin repurposing (18). Second, participants with AD eligible for trial participation are typically at an age where other age-related pathology is likely to be present and quantifiable at baseline. AD has been associated with increased prevalence or accelerated progression of pathology in organ systems relevant to our previously proposed imaging battery (9), including cardiovascular dysfunction (20,21), periodontal disease (22,23), and retinal neurodegeneration (24).

While AD-specific outcomes, including results from CNS imaging, have been reported in a separate publication (25), we here present completion rates, participant burden, and technical challenges, along with exploratory results on the effects of six months of intermittent rapamycin treatment on imaging markers of age-related pathology in the heart, skeleton, body musculature, retina, periodontium, and vasculature.

## 2. Methods

### 2.1 Study design, Participants, and Regulatory Approval

The study was conducted as a single-center, open-label, single-arm, pilot trial at Theme Aging and Inflammation, Medical Unit Aging, Memory Clinic, Karolinska University Hospital, Stockholm, Sweden, between September 2023 and January 2025. Participants were recruited from the Memory Clinic patient population. Eligible participants were aged 50 to 80 years, had a clinical diagnosis of mild cognitive impairment (MCI) or mild dementia of the Alzheimer’s type according to NIA-AA 2018 criteria (26), and were amyloid-positive as confirmed by cerebrospinal fluid (CSF) sample. For participants with dementia, the disease had to be in an early stage, defined as a Montreal Cognitive Assessment (MoCA) score ≥18. Exclusion criteria included major diseases that could interfere with safe engagement in the trial, presence of contraindications to rapamycin, significant obesity, untreated hyperlipidemia, and recent use of immunosuppressive or experimental AD medications. Full eligibility criteria are detailed in the previously published study protocol (19).

The study was approved by the Swedish Medical Products Agency (5.1-2023-8283) and the Swedish Ethical Review Authority (2023-03075-02; 2023-00611-01). The study was conducted in accordance with the Declaration of Helsinki and ICH Good Clinical Practice guidelines. Written informed consent was obtained from all participants and a study partner prior to enrolment. The trial was registered at ClinicalTrials.gov (NCT06022068) and EudraCT (2023-000127-36).

### 2.2 Intervention

Participants received oral rapamycin (sirolimus; Rapamune, Pfizer) 7 mg once weekly for 26 weeks. An intermittent dosing regimen was chosen to preferentially inhibit mTORC1, which has been hypothesized to mediate the beneficial geroprotective effects of rapamycin, while minimizing inhibition of mTORC2, thought to be associated with potential adverse metabolic effects (11). Similar intermittent regimens have been used in other geroprotection trials targeting mTOR (27,28).

### 2.3 Imaging Assessments

Imaging was performed at baseline (within six weeks before first dose) and at end-of-treatment (between two weeks prior and four weeks following last dose (29)). Brain imaging outcomes, including [^18^F]FDG PET, brain MRI, and CSF biomarkers, as well as pharmacokinetic analysis of drug concentration in blood have been reported in separate publications (25,29). Here we describe the imaging battery assessing age-related pathology outside the central nervous system.

#### 2.3.1 Retinal optical coherence tomography (OCT)

Retinal images were obtained with a Zeiss Cirrus HD-OCT (Version 8.0). The retinal ganglion cell and inner plexiform layer (GC+IPL) thickness (µm) was measured using the macular cube 512×128 protocol and the peripapillary retinal nerve fiber layer (pRNFL) thickness (µm) was measured using the optic disc cube 200×200 protocol. OCT scans with poor quality (e.g., poor centration, blurred images, presence of blink, motion artifacts or poor fixation) were excluded. Thickness measurements from the macular region was presented as average GC+IPL, and the pRNFL as a total average and in quadrants (superior, inferior, nasal and temporal).

#### 2.3.2 Quantitative computed tomography: bone mineral density and body composition

Bone mineral density and paraspinal muscle composition were assessed using quantitative CT (qCT) acquired during the [^18^F]FDG PET/CT session on a GE Discovery MI 5 PET/CT system. Helical CT images were acquired with standard reconstruction kernel at 1.25 mm slice thickness, covering the L1 to L2 vertebral region. A Mindways QCT calibration phantom containing inserts of known density (0, 50, 100, and 200 mg/cm^3^ hydroxyapatite equivalent) cylinders was positioned beneath the participant for all scans.

Trabecular volumetric bone mineral density (vBMD, mg/cm^3^) at L1 and L2 was extracted using a fully automated pipeline: Vertebral bodies were segmented using TotalSegmentator (v2) (30), isolated from posterior elements, eroded to exclude cortical bone, and calibrated against the phantom using a per-scan linear regression (R^2^ > 0.997 in all scans). Paraspinal (erector spinae) muscle composition at L1 to L2 was quantified within a fascial envelope derived from TotalSegmentator muscle labels. Skeletal muscle density (SMD), low-attenuation muscle fraction, intermuscular adipose tissue (IMAT), and muscle cross-sectional area (CSA) were determined by HU-based tissue classification after scanner drift correction. Full methodological details, including the vertebra identification algorithm, tissue classification thresholds, and IMAT validation, are provided in Supplementary Methods S1.

#### 2.3.3 Periodontal imaging

Periodontal imaging status was assessed using CT images from the PET/CT session and from high-resolution T2-weighted Short Tau Inversion Recovery (STIR) MR of the upper jaw (maxilla), acquired on a GE Signa Premier 3T scanner. The imaging field of view included teeth and surrounding periodontal tissues (i.e., the periodontium). Each tooth was assessed by a rater (neuroradiologist with extensive expertise in dentomaxillofacial imaging / neuroradiologist) as healthy, unhealthy (presence of periodontal oedema), or not rateable. The rater was blinded to timepoint, clinical and demographic information. Per-tooth ratings were summarized descriptively as counts of healthy/unhealthy teeth at each timepoint.

#### 2.3.4 Cardiac MRI and CT

Cardiac MRI was performed on a 1.5T MAGNETOM Sola scanner (Siemens Healthineers, Forchheim, Germany). Patients were instructed to not consume caffeine 24h prior to the examination. The protocol included full coverage cine imaging, T1 mapping, T2 mapping, late gadolinium enhancement (LGE) imaging, extra cellular volume (ECV) mapping, 4D Flow MRI and quantitative rest and stress perfusion imaging.

Rest perfusion images were collected before pharmacological vasodilation (stress) was induced by a 10 seconds injection of regadenoson (400 µg/5mlL Rapiscan, GE healthcare AB) to assess myocardial stress perfusion and perfusion reserve. Quantitative perfusion maps were acquired as three short-axis slices covering the left ventricle and three long-axis slices (2, 3, 4 chamber view). During image acquisition, gadobutrol (0.05 mmol/kg, Gadovist, Bayer AB, Solna, Sweden) was administered. Quantitative perfusion was obtained using a dual-sequence, single bolus technique as previously described (31). Perfusion reserve was calculated as the ratio of stress to rest perfusion.

A total of gadobutrol of 0.15 mmol/kg was administered intravenously for calculation of ECV, calibrated to blood haematocrit and for determining the presence of LGE.

Outcomes included: ejection fraction, global longitudinal strain, peak filling rate, peak ejection rate, ventricular volumes, ventricular mass, ECV, stress perfusion, myocardial perfusion reserve, cardiac output, and heart rate.

CMR images were analyzed by one level 3 EACVI certified physician and one level 3 SCMR certified CMR technologist using Syngo (Siemens Healthineers, Forchheim, Germany) and Segment (version 4.1.0.3, Medviso AB, Lund, Sweden) (32). Assessment of 4D Flow MRI datasets were separately analysed using dedicated research software (MASS; Leiden University Medical Center, Leiden, The Netherlands).

Epicardial adipose tissue (EAT) volume was quantified from the CT component of the PET/CT (see below). A pericardial-proxy envelope was constructed from the per-slice convex hull of the cardiac cluster (myocardium, chambers, proximal great vessels), dilated 8 mm, and anatomically constrained. Fat voxels were identified using Hounsfield unit thresholds [−190, −30] (See Supplementary Methods S2).

#### 2.3.5 Aortic pulse wave velocity MRI

Aortic stiffness was assessed using phase-contrast MRI, acquired during the cardiac MRI session. Through-plane velocity-encoded images were obtained at the ascending and descending aorta. Pulse wave velocity (m/s) was calculated by measuring the distance between the two measurement points and determining the time difference between the onset of the flow waveform between the two locations and dividing these measures.

#### 2.3.6 [^18^F]FDG PET

In addition to brain [^18^F]FDG PET reported separately (25), the PET/CT acquisition provided data for exploratory analyses of metabolic activity in three peripheral tissues: the optic nerve head (ONH), periodontal soft tissues, and the descending thoracic aorta wall. All participants underwent [^18^F]FDG PET/CT on a GE Discovery MI 5 scanner at baseline and follow-up (see Supplementary Methods S3).

For all PET analyses, the primary quantitative metric was the fractional uptake rate (FUR), a proxy for the net influx rate constant (*K*_i_). FUR was computed as tissue activity concentration divided by the area under the plasma time-activity curve (AUC) from injection to the scan midpoint. The input function was constructed by combining an image-derived input function from the descending aorta with manual venous plasma samples (33). FUR was selected over the commonly used standardized uptake value (SUV) because it accounts for individual differences in plasma tracer kinetics, providing a more physiologically informative measure of tissue glucose metabolism. SUV results, which showed consistent findings, are reported in Supplementary Tables.

##### Optic nerve head

Regions of interest over the left and right ONH were manually delineated on the reconstructed PET images under blinded conditions. Because the ONH is substantially smaller than the PET spatial resolution (∼5 mm FWHM), we employed resolution-robust intensity measures: the mean of the 150 highest-intensity voxels within each mask (corresponding to approximately two resolution elements) was selected as the primary metric. Detailed rationale and additional metrics are provided in Supplementary Methods S4.

##### Periodontal soft tissues

Individual teeth were segmented from CT images using TotalSegmentator (v2, teeth task). A 4 mm periodontal shell ROI was generated around each tooth, with the tongue (dilated 3 mm) subtracted to minimize contamination. ROIs were transformed from CT to PET space via rigid registration (ANTsPy). Analysis was restricted to the upper jaw, and only teeth present at both timepoints were included (mean 14.2 teeth per subject). Jaw-level metrics were obtained by averaging FUR mean and 90th percentile (p90) across all qualifying upper jaw teeth per session. Full details are provided in Supplementary Methods S5.

##### Descending thoracic aorta

A vessel wall ROI was constructed by isolating the descending thoracic aorta between vertebral levels T4 and T12 using TotalSegmentator (v2) and applying a 2D per-slice binary erosion to generate an inner-ring shell representing the vessel wall. The superior vena cava (SVC), eroded to minimize partial-volume effects, served as the blood pool reference for tissue-to-background ratio (TBR) calculation. FUR_mean_ and TBR_mean_ (SUV_mean,wall_ / SUV_mean,blood pool_) were computed for the vessel wall ROI. Full details are provided in Supplementary Methods S6.

### 2.4 Feasibility assessment

The primary aim of this study was to assess the feasibility of deploying a multi-modal imaging battery to evaluate geroprotective interventions. Feasibility was evaluated based on three criteria: 1) completion rates for each imaging modality, defined as the proportion of participants who completed both baseline and follow-up assessments; 2) time burden, reported as approximate duration of each imaging procedure; and 3) protocol deviations or technical challenges encountered during data acquisition or analysis, reported descriptively.

### 2.5 Statistical analysis

For continuous imaging outcomes, pre-post comparisons were performed using two-sided paired t-tests. Effect sizes are reported as Cohen’s d_z_ (mean change divided by the standard deviation of change). Periodontal MRI data were reported descriptively as per-tooth counts of healthy and unhealthy ratings at each timepoint; this outcome was compared using a Wilcoxon signed-rank test. Given the exploratory nature of this pilot study, no correction for multiple comparisons was applied. All analyses were performed in Python (SciPy) and R version 4.2.1.

## 3. Results

### 3.1 Participants

Fourteen participants were enrolled in the study. One participant discontinued within two weeks of initiating treatment due to intolerance of the study drug (nausea) and did not complete follow-up imaging, resulting in 13 participants with complete imaging data. Baseline demographic and clinical characteristics are presented in Table 1. Two participants (15%) initially received reduced rapamycin doses (2 mg and 4 mg weekly) due to safety considerations or side effects and were escalated to higher doses (4 mg and 7 mg weekly, respectively) after the 13-week follow-up. None of the participants were prescribed bisphosphonates. Full dosing details, adherence data, and adverse events are reported elsewhere (25).

**Table 1.** Demographics and clinical characteristics of study participants (n = 14).

| Characteristic | Value |
| --- | --- |
| **Demographics** |  |
| Age, years, mean ± SD (range) | 60.6 ± 4.3 (51-67) |
| Female / Male, n | 8 / 6 |
| BMI, kg/m<sup>2</sup>, mean ± SD (range) | 25.4 ± 5.1 (18.7-33.2) |
| **Diagnosis** |  |
| MCI due to AD, n (%) | 6 (43) |
| Mild dementia of Alzheimer's type, n (%) | 8 (57) |
| **Cognitive Function** |  |
| MoCA score, mean ± SD (range) | 23.8 ± 3.1 (19-28) |
| **Cardiovascular Risk Factors** |  |
| Hypertension, n (%) | 3 (21) |
| Hyperlipidemia, n (%) | 3 (21) |
| Current smoker, n (%) | 1 (7) |
| **Metabolic Parameters** |  |
| HbA1c, mmol/mol, mean ± SD | 36.1 ± 3.2 |
| Fasting glucose, mmol/L, mean ± SD | 5.4 ± 0.4 |
| **Genetic Profile** |  |
| ApoE ε4 carrier, n (%) | 11 (79) |
| **Concomitant Medications** |  |
| Cholinesterase inhibitor, n (%) | 14 (100) |
| Antihypertensive, n (%) | 4 (29) |
| Lipid-lowering agent, n (%) | 5 (36) |
| Antidepressant, n (%) | 6 (43) |
| Vitamin D supplement, n (%) | 2 (14) |
| Calcium supplement, n (%) | 1 (7) |

### 3.2 Feasibility of the imaging battery

The multi-modal imaging battery demonstrated high feasibility for most modalities in this patient population. All 13 participants who completed the study successfully underwent both baseline and follow-up retinal OCT, [^18^F]FDG PET/CT, quantitative CT bone density and body composition measurements, and periodontal imaging, yielding 100% completion rates for these modalities (except for one incomplete OCT macular scan for the right eye for one participant follow-up). In contrast, cardiac MRI and aortic pulse wave velocity assessments were completed in only 9 of 13 participants (69%), with the reduced completion rate attributable to a healthcare worker strike that limited scanner access during the study period rather than participant-related factors. Out of these 9, myocardial perfusion imaging was successfully acquired in 6 participants. One participant had consumed caffeine-containing beverages in too close proximity to the examination, and for two participants suboptimal hyperemia precluded reliable perfusion estimates.

All imaging procedures were well-tolerated, with no adverse events attributable to the imaging assessments. The total imaging burden per participant was approximately 12 hours across all modalities, distributed over six separate visits during the study. No participants withdrew from the study due to imaging-related discomfort or inconvenience, suggesting acceptable participant burden for this type of assessment battery.

### 3.3 Imaging outcomes

A summary of all imaging outcomes is presented in Table 2. For most imaging parameters, no statistically significant changes were observed between baseline and follow-up assessments. A set of outcomes showed nominally significant changes (p < 0.05, uncorrected for multiple comparisons), as detailed below.

**Table 2.** Imaging outcomes at baseline and follow-up.

| Organ system | Outcome | n | Baseline | Follow-up | $\Delta$ (95% CI) | $d_z$ | p |
| --- | --- | --- | --- | --- | --- | --- | --- |
| **Retina** |  |  |  |  |  |  |  |
| | GCL, left eye ($\mu\text{m}$) | 13 | $80.3 \pm 4.6$ | $81.0 \pm 4.1$ | 0.69 (0.02, 1.36) | 0.62 | 0.044 |
| | GCL, right eye ($\mu\text{m}$) | 12* | $80.9 \pm 4.1$ | $81.2 \pm 4.1$ | 0.25 (-0.36, 0.86) | 0.26 | 0.389 |
| | RNFL, left eye ($\mu\text{m}$) | 13 | $92.5 \pm 11.0$ | $91.8 \pm 9.6$ | -0.66 (-2.19, 0.86) | -0.26 | 0.361 |
| | RNFL, right eye ($\mu\text{m}$) | 13 | $92.8 \pm 9.6$ | $93.4 \pm 9.4$ | 0.63 (-0.87, 2.14) | 0.25 | 0.377 |
| | ONH $\text{FUR}_{\text{top150}}$, bilateral ($\times 10^{-3} \text{ min}^{-1}$) | 13 | $13.0 \pm 2.0$ | $14.1 \pm 2.3$ | 1.1 (0.0, 2.1) | 0.63 | 0.040 |
| | ONH $\text{FUR}_{\text{top150}}$, left ($\times 10^{-3} \text{ min}^{-1}$) | 13 | $13.0 \pm 1.9$ | $14.1 \pm 2.4$ | 1.1 (0.1, 2.1) | 0.66 | 0.030 |
| | ONH $\text{FUR}_{\text{top150}}$, right ($\times 10^{-3} \text{ min}^{-1}$) | 13 | $13.0 \pm 2.1$ | $14.1 \pm 2.4$ | 1.0 (-0.2, 2.3) | 0.52 | 0.080 |
| **Bone & Muscle** |  |  |  |  |  |  |  |
| | L1-L2 vBMD ($\text{mg}/\text{cm}^3$) | 13 | $111.9 \pm 26.1$ | $114.1 \pm 27.9$ | 2.11 (-2.19, 6.41) | 0.30 | 0.307 |
| | Muscle SMD (HU) | 13 | $40.7 \pm 8.1$ | $40.4 \pm 8.6$ | -0.34 (-2.44, 1.75) | -0.10 | 0.729 |
| | Muscle CSA ($\text{cm}^2$) | 13 | $33.2 \pm 9.0$ | $32.2 \pm 9.7$ | -0.97 (-1.98, 0.04) | -0.58 | 0.058 |
| | IMAT (%) | 13 | $3.4 \pm 2.2$ | $3.7 \pm 2.8$ | 0.33 (-0.21, 0.86) | 0.37 | 0.209 |
| **Periodontium** |  |  |  |  |  |  |  |
| Periodontium | Teeth with oedema (MRI) | 13 | $1.2 \pm 1.9$ | $1.2 \pm 1.5$ | 0.00 (-0.49, 0.49) | 0.00 | >0.99 |
| | Peridental $\text{FUR}$ mean ($\times 10^{-3} \text{ min}^{-1}$) | 13 | $9.39 \pm 1.43$ | $9.78 \pm 1.24$ | 0.39 (-0.20, 0.98) | 0.40 | 0.177 |
| | Peridental FUR p90 ($\times 10^{-3}$ min<sup>-1</sup>) | 13 | 14.46 ± 2.38 | 15.15 ± 2.16 | 0.69 (-0.25, 1.63) | 0.44 | 0.135 |
| **Heart** |  |  |  |  |  |  |  |
| | Heart rate (bpm) | 9 | 69.7 ± 13.1 | 75.7 ± 15.5 | 5.96 (-4.59, 16.51) | 0.43 | 0.229 |
| | Cardiac output (L/min) | 8** | 4.7 ± 0.5 | 5.7 ± 1.1 | 1.03 (0.25, 1.81) | 1.10 | 0.017 |
| | LV mass (g) | 9 | 99.1 ± 29.7 | 102.4 ± 31.4 | 3.39 (0.09, 6.69) | 0.79 | 0.046 |
| | LV ejection fraction (%) | 9 | 64.8 ± 4.2 | 64.7 ± 6.4 | -0.03 (-5.29, 5.23) | -0.00 | 0.989 |
| | Global long. strain (%) | 9 | -21.9 ± 1.4 | -21.3 ± 2.3 | 0.63 (-0.73, 1.99) | 0.36 | 0.316 |
| | ECV (%) | 8** | 25.8 ± 2.9 | 27.0 ± 3.9 | 1.25 (-0.74, 3.24) | 0.53 | 0.180 |
| | Perfusion reserve | 6*** | 2.5 ± 0.5 | 2.9 ± 0.4 | 0.38 (-0.45, 1.22) | 0.48 | 0.290 |
| | E-wave KE, average (μJ/mL) | 9 | 6.5 ± 2.2 | 7.0 ± 2.9 | 0.53 (-1.80, 2.86) | 0.21 | 0.613 |
| | E-wave KE, peak (μJ/mL) | 9 | 15.0 ± 4.5 | 15.7 ± 6.1 | 0.64 (-4.37, 5.65) | 0.12 | 0.776 |
| | A-wave KE, average (μJ/mL) | 9 | 9.4 ± 4.0 | 11.8 ± 5.0 | 2.41 (0.08, 4.74) | 0.53 | 0.044 |
| | A-wave KE, peak (μJ/mL) | 9 | 17.1 ± 5.2 | 19.7 ± 5.5 | 2.59 (0.45, 4.73) | 0.48 | 0.024 |
| **Vasculature** |  |  |  |  |  |  |  |
| | PWV (m/s) | 9 | 7.8 ± 2.0 | 7.5 ± 1.9 | -0.28 (-2.02, 1.46) | -0.12 | 0.719 |
| | Aortic wall FUR mean ($\times 10^{-3}$ min<sup>-1</sup>) | 13 | 8.1 ± 1.1 | 8.4 ± 1.3 | 0.3 (-0.5, 1.1) | 0.21 | 0.460 |
| | Aortic wall TBR mean | 13 | 1.14 ± 0.11 | 1.18 ± 0.14 | 0.04 (-0.02, 0.11) | 0.39 | 0.190 |
| | EAT volume (mL) | 13 | 90.2 ± 45.8 | 94.2 ± 45.3 | 3.9 (-3.8, 11.6) | 0.31 | 0.290 |

Values are mean ± SD. $\Delta$ = mean paired difference (follow-up minus baseline) with 95% confidence interval. $d_z$ = Cohen's $d_z$ . GCL = ganglion cell layer; RNFL = retinal nerve fiber layer; ONH = optic nerve head; FUR = fractional uptake rate; vBMD = volumetric bone mineral density; SMD = skeletal muscle density; CSA = cross-sectional area; IMAT = intermuscular adipose tissue; LV = left ventricle; ECV = extracellular volume; E-wave = early diastolic filling; A-wave = atrial contribution to late diastolic filling; KE = kinetic energy; PWV = pulse wave velocity; TBR = tissue-to-background ratio; EAT = epicardial adipose tissue. All p-values from two-sided paired t-tests except for teeth with oedema (Wilcoxon signed-rank test), uncorrected for multiple comparisons.
\*\*For cardiac output, one baseline value was unavailable due to technical issues. For ECV, one follow-up value was unavailable due to a technical acquisition error.
\*\*\*For myocardial perfusion one participant had consumed caffeine-containing beverages in too close proximity to the examination, and for two participants suboptimal hyperemia precluded reliable perfusion estimates.

#### 3.3.1 Retina

All 26 eyes from 13 patients underwent successful OCT examinations of both the macula and optic disc at visit one. At the follow-up visit, one macular scan of a right eye was incomplete, and did not generate any data. Average RNFL thickness did not change significantly in either eye (left: p = 0.361; right: p = 0.377). GCL thickness showed a nominally significant increase in the left eye (p = 0.044) but not in the right eye (p = 0.389). Quadrant-specific RNFL measurements are provided in Supplementary Table S1. Increased age was unexpectedly correlated with a thicker GC_IPL (r = 0.62; p = 0.02) and pRNFL (r = 0.69; p = 0.009).

[^18^F]FDG uptake at the ONH, quantified as FUR_top150_ (the mean fractional uptake rate across the 150 highest-intensity voxels), increased from baseline to follow-up. The bilateral average showed a nominally significant increase of 8.2% (p = 0.040), with the effect reaching significance in the left eye (p = 0.030) but not in the right eye (p = 0.080). The direction was consistent across all quantification methods (Supplementary Tables S1 and S4).

**Figure 1.**
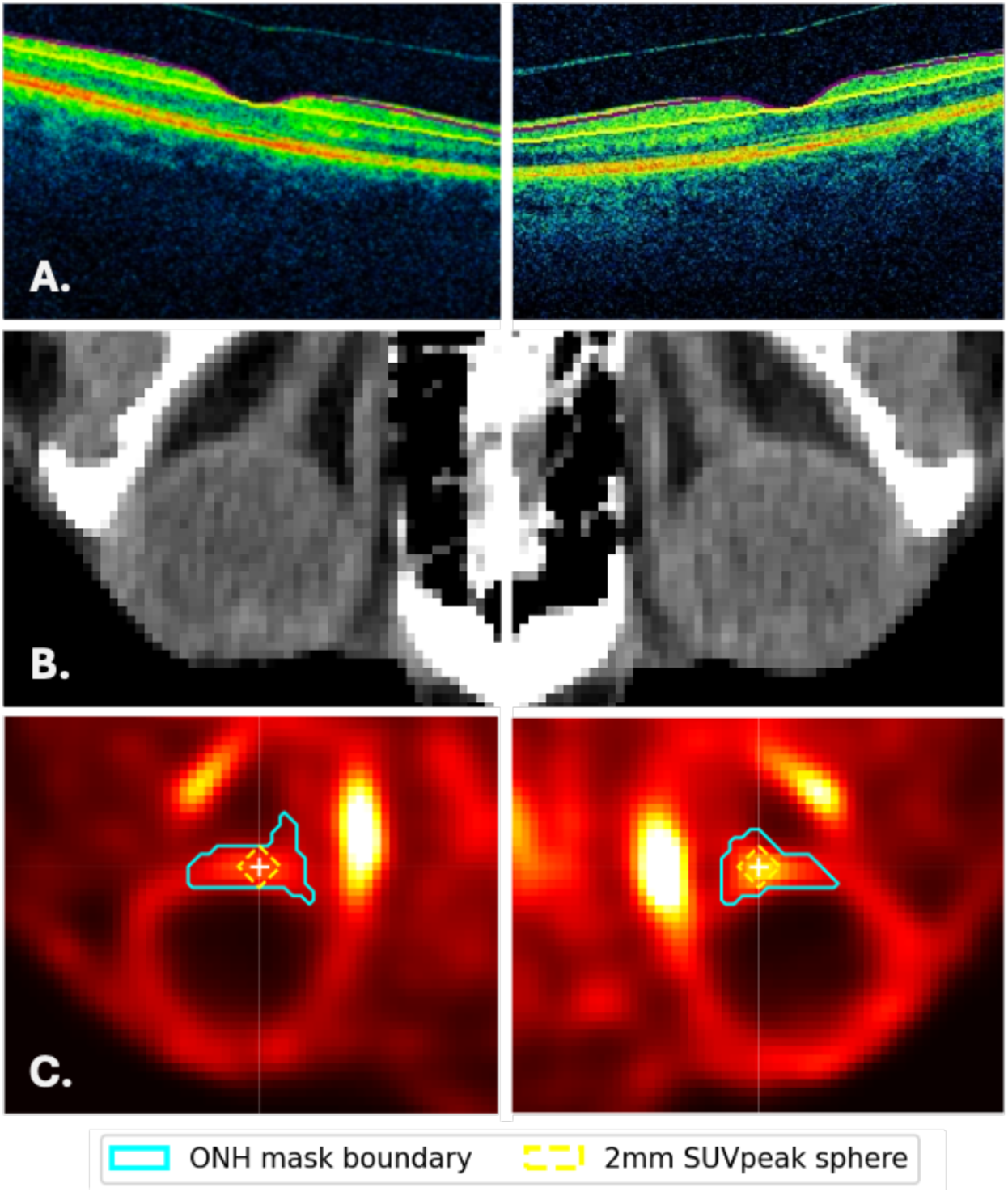
Retinal and optic nerve head imaging from a representative participant. (A) Horizontal macular OCT B-scans (macular cube 512×128) for the right and left eye with purple and yellow line marking the GC+IPL layer. (B) Axial CT images at the level of the orbits showing the optic nerves bilaterally on which ONH masks were drawn. (C) Corresponding [^18^F]FDG PET images (Bq/mL) with the search mask boundary (solid line) and the 2 mm radius FUR peak sphere (dashed line) centered on the maximum-intensity voxel.

#### 3.3.2 Bone and muscle composition

Trabecular vBMD at L1 to L2 did not change significantly (p = 0.307). L1 and L2 individually showed similar non-significant trends (Supplementary Table S1). Skeletal muscle density and intermuscular adipose tissue fraction remained stable. Muscle cross-sectional area showed a numerical decrease (p = 0.058). Additional bone and muscle outcomes are provided in Supplementary Table S1.

**Figure 2.**
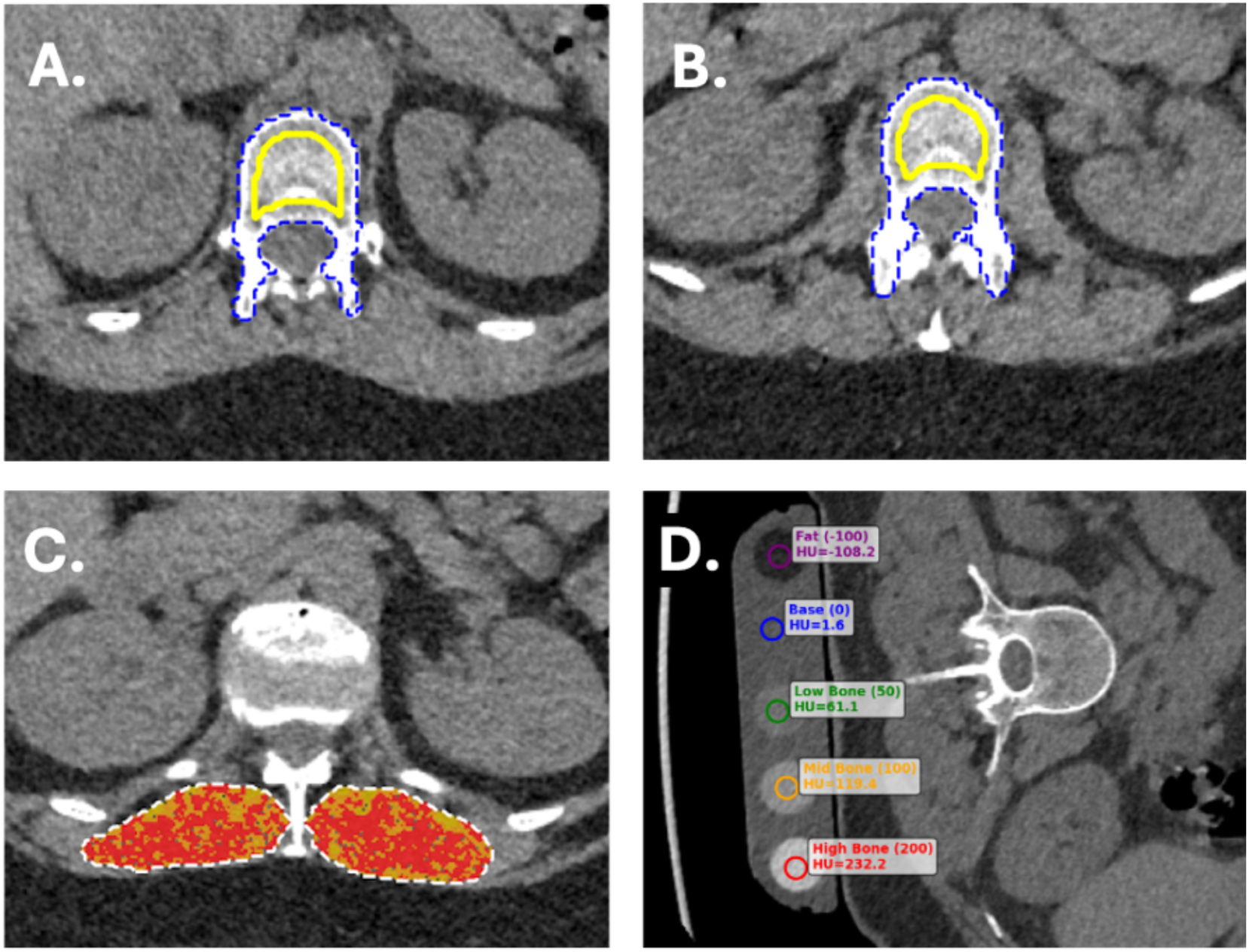
CT-derived bone and muscle assessment from a representative participant. (A, B) Axial CT images at the L1 and L2 vertebral levels, respectively, showing the full vertebral body segmentation mask (outline) and the eroded trabecular region of interest used for volumetric bone mineral density estimation. (C) Erector spinae muscle composition within the fascial envelope at L1 to L2: muscle tissue (red), low-attenuation muscle (orange), and intermuscular adipose tissue (IMAT, yellow). (D) QCT calibration phantom with cylinder masks (colored outlines) over the known-density inserts (0, 50, 100, and 200 mg/cm³ hydroxyapatite equivalent) used to derive the per-scan calibration equation.

#### 3.3.3 Periodontium

The mean number of teeth with signs of periodontal oedema on MRI remained stable, with no significant change observed over time (Wilcoxon signed-rank p > 0.99). At the individual level, 2 participants showed improvement (i.e., fewer teeth with oedema), 3 showed worsening (i.e., more teeth with oedema), and 8 showed no change.

Peridental [^18^F]FDG uptake, assessed at the jaw level, showed numerical increases from baseline to follow-up for both FUR mean (+4.2%) and FUR p90 (+4.8%), but neither showed significant change (p = 0.177 and p = 0.135, respectively).

**Figure 3.**
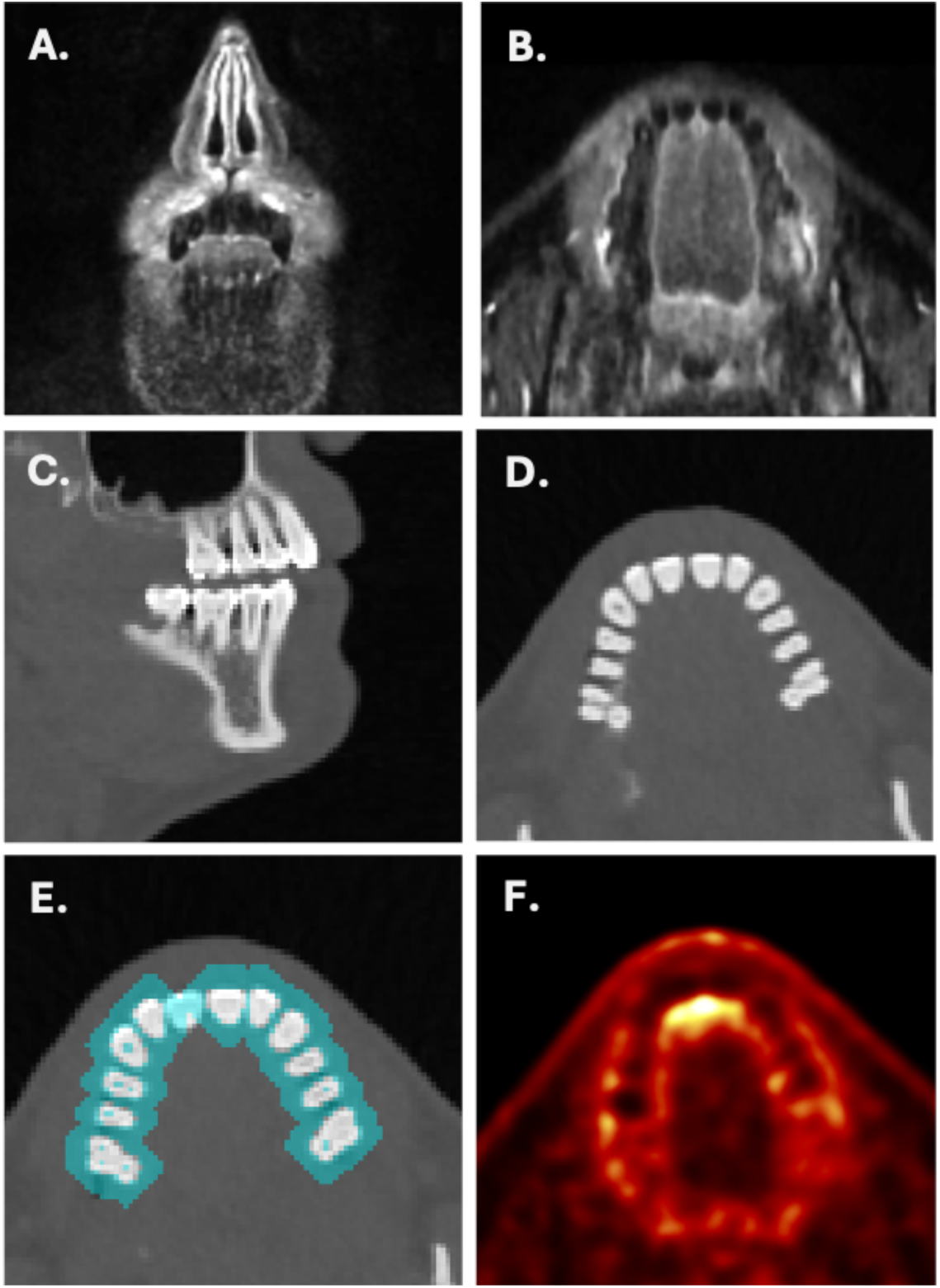
Periodontal imaging from a representative participant. (A, B) Coronal and axial T2-weighted STIR MRI of the upper jaw and (C, D) sagittal and axial CT images of the dentition used for per-tooth assessment of periodontal bone loss. (E) Axial CT with peridental shell-shaped regions of interest (cyan) generated by dilating each tooth segmentation mask by 4 mm and subtracting the original tooth volume. These masks were then applied to the (F) corresponding [^18^F]FDG PET image to extract average uptake in the peridental soft tissues.

#### 3.3.4 Heart

Among cardiac MRI parameters, two outcomes showed nominally significant changes. Cardiac output, available for 8 participants, increased (p = 0.017). Left ventricular mass, available for 9 participants, also increased (p = 0.046). Left ventricular ejection fraction (p = 0.989), global longitudinal strain (p = 0.316), and extracellular volume fraction (p = 0.180; available for 8 participants) showed no significant changes. Myocardial perfusion reserve, available for 6 participants, showed a numerical increase but no statistically significant change (p = 0.290). Additional cardiac parameters including ventricular volumes are provided in Supplementary Table S1.

4D Flow analysis of diastolic kinetic energy was available for 9 participants. Early diastolic (E-wave) kinetic energy showed no significant change (average: p = 0.613; peak: p = 0.776). In contrast, late diastolic (A-wave) kinetic energy, reflecting atrial contribution to ventricular filling, showed nominally significant increases for both average (p = 0.044) and peak (p = 0.024) A-wave kinetic energy. Additional 4D Flow parameters including ventricular washout fractions are provided in Supplementary Table S1.

**Figure 4.**
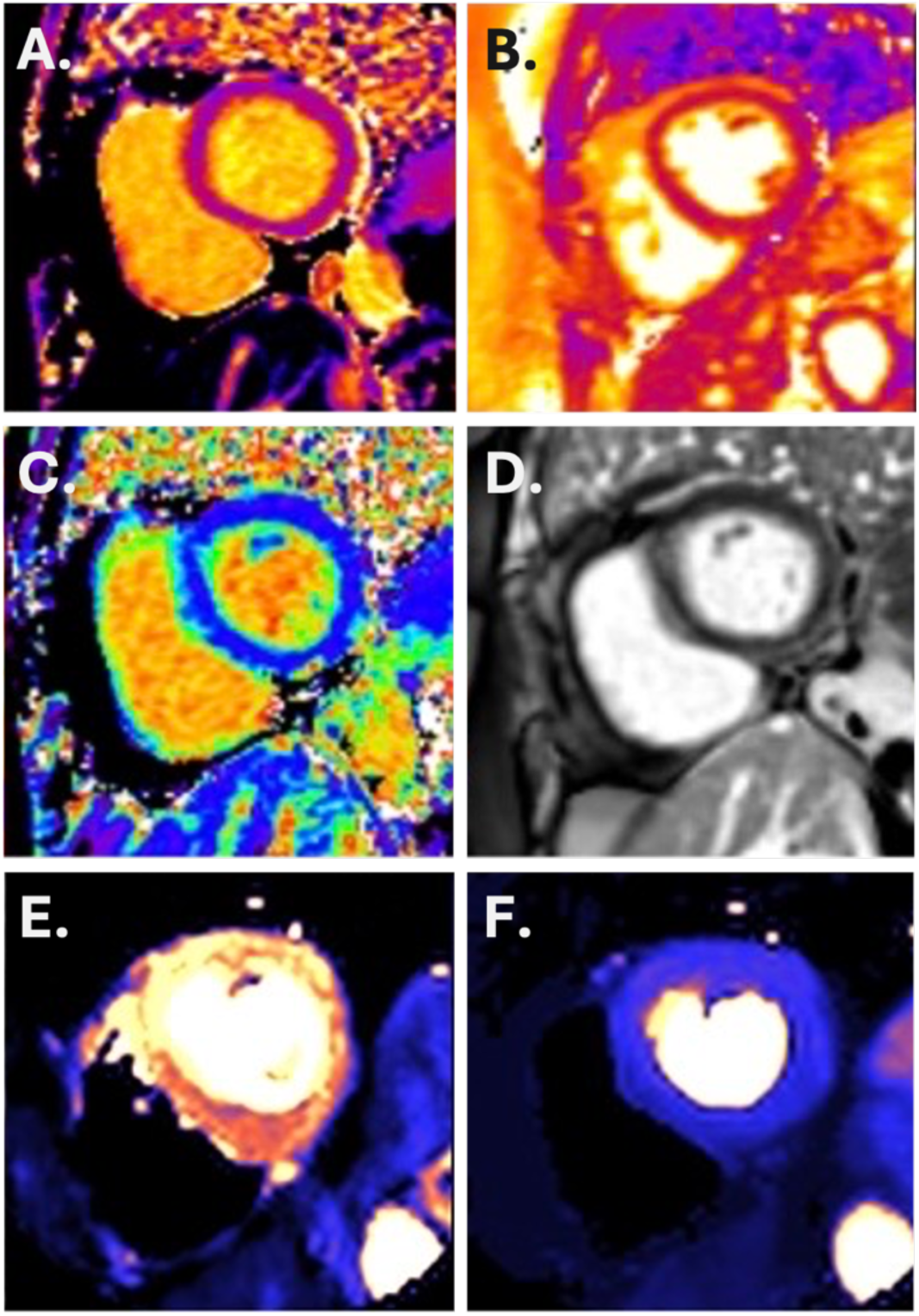
Cardiac MRI short-axis views from a representative participant. (A) Native T1 map (ms). (B) Native T2 map (ms). (C) Extracellular volume fraction (ECV, %). (D) Late gadolinium enhancement (LGE) image. (E) Quantitative stress perfusion map (mL/min/g) acquired during regadenoson-induced vasodilation. (F) Quantitative rest perfusion map (mL/min/g).

#### 3.3.5 Vasculature and epicardial adipose tissue

Aortic pulse wave velocity did not change significantly (p = 0.719). Aortic vessel wall [^18^F]FDG uptake, assessed as FUR_mean_ (p = 0.460) and TBR_mean_ (p = 0.190), showed no significant changes. Blood pool SUV_mean_ in the SVC was stable between sessions (p = 0.90), confirming its suitability as a reference region. Epicardial adipose tissue volume did not change significantly (p = 0.290).

**Figure 5.**
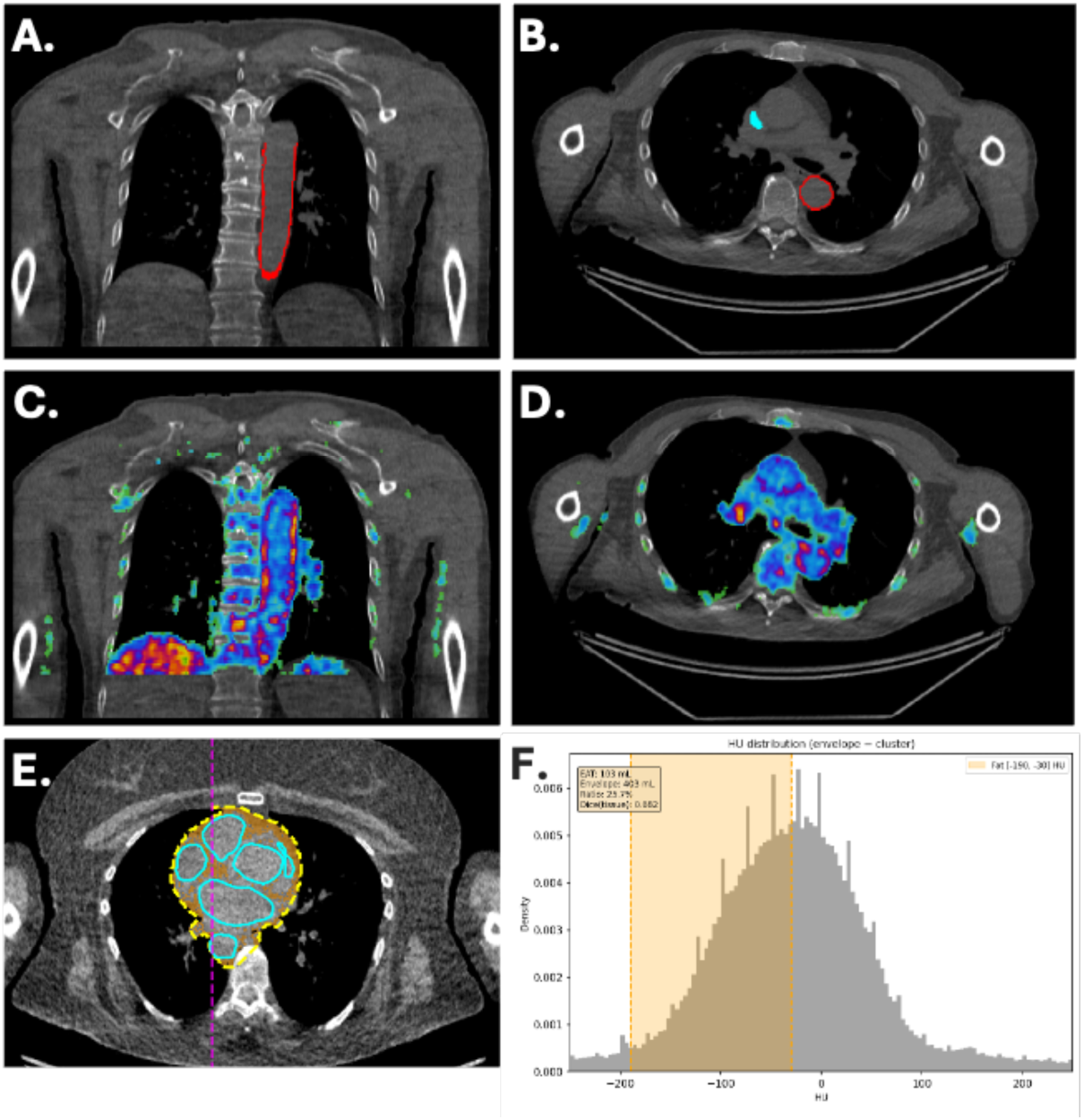
Vascular [^18^F]FDG uptake and epicardial adipose tissue quantification from a representative participant. (A) Coronal CT image showing the descending thoracic aorta vessel wall region of interest (red outline), constructed by 2D per-slice erosion of the aorta cross-section between vertebral levels T4 and T12. (B) Axial CT at the same level showing the aortic wall ROI and the superior vena cava blood pool reference region (cyan), used for tissue-to-background ratio calculation. (C, D) Corresponding coronal and axial views with [^18^F]FDG PET uptake overlaid. (E) Axial CT showing the pericardial-proxy envelope used for epicardial adipose tissue (EAT) quantification, with classified EAT voxels in orange. (F) Hounsfield unit distribution within the envelope, with the EAT classification range (−190 to −30 HU) highlighted.

### 3.4 Exposure-response analysis

Exploratory analyses examined correlations between rapamycin blood concentration (measured 48 hours post-dose at week 13) and change in imaging outcomes. Two participants increased their dose of rapamycin (from 2 / 4 mg to 4 / 7 mg, respectively) after this timepoint and have therefore been excluded from this analysis (n=11). For bone mineral density, a positive but non-significant correlation was observed (r = 0.41, p = 0.211). Among CT-derived muscle composition metrics, higher drug concentration was significantly associated with increased skeletal muscle density (r = 0.64, p = 0.035) and (albeit non-significant) reduced low-attenuation muscle proportion (r = −0.59, p = 0.058) and attenuated loss of muscle cross-sectional area (r = −0.53, p = 0.097). Full correlation results are reported in Supplementary Table S2.

## 4. Discussion

This pilot study demonstrates that a multi-modal, multi-organ *in vivo* imaging battery can be effectively deployed within a clinical trial setting to assess candidate geroprotective interventions. Completion rates were high across all imaging modalities, and participant burden was acceptable. We examined the effects of six months of intermittent rapamycin treatment on imaging markers of age-related pathology in the retina, skeleton, periodontium, skeletal musculature, heart, and vasculature. For most outcomes, no statistically significant changes were observed between baseline and follow-up. Several outcomes showed nominally significant increases, including cardiac output, left ventricular mass, A-wave diastolic kinetic energy, left GCL thickness, and optic nerve head [^18^F]FDG uptake, but these findings were not corrected for multiple comparisons and should be interpreted with caution.

To our knowledge, this is the first practical implementation of an imaging-based approach for evaluating GPIs. All imaging modalities included in the battery are drawn from routine clinical practice and were successfully integrated into the trial infrastructure without major protocol deviations or technical failures. The [^18^F]FDG PET/CT session proved particularly versatile, enabling quantification of metabolic activity across multiple peripheral tissues (optic nerve head, periodontium, aortic vessel wall) as well as bone mineral density, muscle composition, and epicardial adipose tissue volume from a single acquisition. This suggests that the approach could serve either as a framework for dedicated geroprotection trials, or as an exploratory add-on to disease-specific trials where geroprotective effects are the secondary outcome. The present study provides practical insights, including considerations around scheduling, participant burden, and data acquisition, that can inform the design of future randomized controlled trials in the field.

Preclinical studies have demonstrated beneficial effects of rapamycin on several of the organ systems assessed in our imaging battery, including cardiac function, retinal neuroprotection, periodontal disease, and bone remodeling (12–16). In the current pilot, we did not observe consistent changes across these domains. Several factors may explain this discrepancy. Most importantly, the single-arm design precludes definitive efficacy conclusions: without a control group, we could only detect reversal of existing pathology, not slowing of progression. Slowing of progression, arguably the more plausible mechanism for a GPI (4,34), would require comparison to an untreated or placebo group. Additional contributing factors include the small sample size, the relatively short treatment duration of six months.

Several outcomes did however reach nominal statistical significance. Two of these relate to the retina: an increase in left GCL thickness and an increase in ONH [^18^F]FDG uptake (35,36). The GCL finding is difficult to interpret in isolation, as retinal thickness typically declines with age; an increase could reflect measurement variability, regression to the mean, or a true biological effect. The [^18^F]FDG increase is similarly exploratory but noteworthy: higher [^18^F]FDG uptake at the optic nerve head could reflect increased metabolic activity in retinal ganglion cell axons, which converge at this location. If confirmed, one possibility is that mTOR inhibition mitigates retinal neurodegenerative processes, consistent with preclinical reports of rapamycin-mediated neuroprotection in ocular and retinal contexts (13). The mean GC-IPL and pRNFL thickness was similar to what expected according to the literature describing age normative data (37). However, the expected age-related decrease in retinal layer thickness was not present in our data. Instead, older age correlated with increased thickness of both GC-IPL and pRNFL thickness. One possibility could be that the younger patients in this small sample had a more severe or advanced neurodegenerative disease stage (e.g., all five participants under the age of 60 were diagnosed with dementia, while only two out eight participants above 60 fulfilled the criteria for dementia).

The cardiac output increase, aligns with preclinical evidence suggesting that rapamycin improves cardiac function in aged mice (35,36). Recently, Moody et al. reported improved diastolic function following 8 weeks of daily rapamycin (1 mg) in six healthy older men, including increased peak filling rate, blood acceleration across the mitral valve, and transmitral blood volume (38). In contrast, cardiac output did not change significantly in their study. Our study observed nominally significant increases in both cardiac output and late diastolic (A-wave) kinetic energy, but not in early diastolic (E-wave) parameters. This difference may reflect population characteristics (our participants had early-stage AD, whereas Moody et al. studied healthy volunteers), dosing regimens (intermittent 7 mg weekly vs. daily 1 mg), or the distinct phases of diastole assessed. Nevertheless, the convergent finding of improved diastolic function across two independent studies with different designs strengthens the hypothesis that mTOR inhibition may benefit cardiac function in aging. Of note, while cardiac output increased significantly in our study, neither heart rate (+6.0 bpm, p = 0.229) nor stroke volume (+2.7 mL, p = 0.564) reached individual significance, suggesting the CO increase reflects a combination of modest changes in both components. The numerically larger contribution from heart rate warrants attention, since elevated resting heart rate can reflect increased sympathetic activation and is generally considered a prognostically unfavorable sign (39). This distinction underscores the importance of larger controlled trials to clarify the mechanism and clinical significance of the cardiac output finding.

The nominally significant increase in left ventricular mass (+3.4 g) was statistically significant (at uncorrected level) but unlikely to be clinically meaningful at this magnitude. Periodontal [^18^F]FDG uptake increased numerically after treatment; however, interpretation is limited because rapamycin commonly causes stomatitis, which can produce transient oral inflammation and elevate local PET signal (40), especially when follow-up imaging is performed shortly after treatment. Conversely, the trend toward decreased muscle cross-sectional area at L1–L2 (−1.0 cm², p = 0.058) warrants consideration in the context of the drug exposure-response analysis. While the group-level change suggests possible muscle atrophy, dose-response correlations suggest that higher rapamycin blood concentration was associated with better preservation of muscle quality: significantly higher muscle density (r = 0.64, p = 0.035), less fatty infiltration (r = −0.59, p = 0.058), and attenuated loss of cross-sectional area (r = −0.53, p = 0.097). These dose-response findings are broadly consistent with the PEARL trial, a 48-week placebo-controlled study of intermittent rapamycin in healthy older adults, which reported significant increases in lean tissue mass for women in the higher-dose group (28). Together, the results suggest that the relationship between rapamycin exposure and skeletal muscle may be more nuanced than a simple catabolic effect, though both studies are limited by small sample sizes and the possibility of skeletal muscle effects should be monitored in future trials.

Vascular [^18^F]FDG uptake in the descending thoracic aorta, a marker of arterial wall inflammation and early atherosclerosis (41), did not change significantly. Similarly, aortic pulse wave velocity and epicardial adipose tissue volume remained stable. These null findings may reflect a true absence of effect. However, the aortic [^18^F]FDG TBR values observed in our cohort (mean 1.14 to 1.18) are consistent with a population with low baseline atherosclerotic burden (42,43), which may have limited the dynamic range for detecting improvement.

While this multi-modal imaging approach proved feasible, it has a set of inherent limitations that should inform future trial design. First, acquisition and analysis of CT, MRI, PET, and OCT data across multiple organ systems is resource-intensive, particularly when repeated at multiple timepoints. Second, imaging-based detection of change requires that some degree of age-related pathology is present at baseline; this constrains the eligible age range and may necessitate enrichment strategies (such as selecting participants with known risk factors) to increase the likelihood of detecting effects. Third, the imaging battery does not capture all relevant age-related processes for geroprotective compounds: some, such as cancer development or immune senescence, cannot be effectively assessed using current medical imaging techniques. Finally, for several processes included in the battery, established non-imaging assessments exist to define the actual disease state (e.g., clinical dental examination for periodontal disease). In these cases, a more accurate disease assessment depends on combining clinical, functional, and image-based assessments. However, when participants are already undergoing imaging for other indications, the marginal burden of adding additional sequences is relatively small (9).

## 5. Conclusion

This study demonstrates that a multi-modal, multi-organ *in vivo* imaging battery is feasible for assessing candidate geroprotective interventions in humans. The approach enables simultaneous evaluation of age-related pathology across different organ systems within a single trial, with high completion rates and acceptable participant burden. The [^18^F]FDG PET/CT session proved particularly efficient, enabling assessment of multiple peripheral tissues and body composition from a single acquisition. While efficacy conclusions cannot be drawn from this uncontrolled pilot, the findings establish a practical framework for future trials. Such trials should employ randomized controlled designs, larger samples, and longer follow-up to provide adequate statistical power. Importantly, detection of slowed progression, rather than reversal of existing pathology, should be considered the primary endpoint, as this more closely reflects the expected mechanism of action of most geroprotective interventions (34). By enabling simultaneous assessment of aging processes across the body within feasible time frames, this imaging-based approach may help bridge the gap between promising preclinical findings and clinical translation of geroprotective interventions.

## Data Availability

Data sharing follows institutional policy. Requests for data access can be directed to the Karolinska Institutet Research Data Office.

## Acknowledgement

We would like to thank Edvin Johansson and Lars Johansson from Antaros Medical for their assistance with the design of MRI and PET imaging protocols. The study was supported by a Longevity Impetus grant from the Norn Group, Åhlén Stiftelsen, Demensfonden, The Swedish Society of Medicine (SLS), Åke Wibergs Stiftelse (M24-0117), Loo and Hans Osterman Stiftelse, Stiftelsen för Ålderssjukdomar Karolinska Institutet, Stiftelsen för Gamla Tjänarinnor, Tore Nilssons Stiftelse för Medicinsk Forskning, Magnus Bergvalls stiftelse, Karolinska Institutet Research Grants, and The Swedish Brain Foundation (PD2024-0444-HK-155).

## Notes

### Competing Interest Statement

The authors have declared no competing interest.

### Clinical Trial

NCT06022068

### Clinical Protocols

https://link.springer.com/article/10.1186/s12883-024-03596-1

### Funding Statement

The study was supported by a Longevity Impetus grant from the Norn Group, Ahlen Stiftelsen, Demensfonden, The Swedish Society of Medicine (SLS), Ake Wibergs Stiftelse (M24-0117), Loo and Hans Osterman Stiftelse, Stiftelsen for Alderssjukdomar Karolinska Institutet, Stiftelsen for Gamla Tjanarinnor, Tore Nilssons Stiftelse for Medicinsk Forskning, Magnus Bergvalls stiftelse, Karolinska Institutet Research Grants, and The Swedish Brain Foundation (PD2024-0444-HK-155).

