## supplementary_materials for "Multi-modal and multi-organ *in vivo* imaging to assess geroprotective interventions in humans: results from a pilot trial of rapamycin in Alzheimer’s Disease"

### Supplementary Methods

#### S1. CT-Derived Bone Mineral Density and Muscle Composition

CT images from the PET/CT session were processed using an automated pipeline. Anatomical segmentation was performed using TotalSegmentator (v2), providing vertebral body and paraspinal muscle labels.

For quantification of bone mineral density (BMD), the L1 and L2 vertebral bodies were identified using a robust detection algorithm (two largest vertebral masks ranked by voxel count, assigned by craniocaudal position). Vertebral bodies were isolated from posterior elements by retaining only the anterior connected component per axial slice and excluding the cranial and caudal 10% (endplates). A 3 mm distance-transform erosion was applied to exclude cortical bone, yielding a trabecular region of interest. A Mindways QCT calibration phantom (inserts of 0, 50, 100, and 200 mg/cm$^3$ hydroxyapatite equivalent) was positioned in the field of view for all scans. Phantom calibration was performed per scan using automatic detection of cylindrical inserts and linear regression of measured HU against known densities (all $R^2 > 0.998$). Trabecular voxel HU values (filtered to −50 to 400 HU) were converted to volumetric bone mineral density (vBMD) in mg/cm$^3$.

Automated L1–L2 trabecular vBMD measurements were validated against independent manual ROI-based measurements performed by an experienced medical physicist (n = 26 scans). The two methods showed excellent agreement ($R^2 = 0.947$, slope = 1.02, bias = +1.6 mg/cm$^3$ [1.4% of the grand mean]; Bland–Altman 95% limits of agreement: −10.4 to +13.6 mg/cm$^3$), confirming that the automated pipeline captures BMD estimates comparably to manual delineation.

For quantification of muscle composition, the left and right autochthonous (erector spinae) muscle masks were combined and a fascial envelope was generated using morphological closing (5 mm spherical structuring element) followed by 2D hole-filling per axial slice, restricted to the L1 to L2 z-range. Within the envelope, voxels were classified by HU after scanner drift correction (estimated from the phantom base insert): muscle tissue (−29 to +150 HU), normal-density muscle (+30 to +150 HU), low-attenuation muscle (−29 to +29 HU), and intermuscular adipose tissue (IMAT; −190 to −30 HU). HU-based IMAT classification was validated against TotalSegmentator's learned intermuscular fat segmentation restricted to the same muscle envelope, yielding a mean Dice coefficient of 0.54 (range 0.06 to 0.75), with Dice values exceeding 0.5 in 73% of sessions.

#### S2. Epicardial Adipose Tissue Quantification

All participants underwent [$^{18}$F]FDG-PET/CT on a GE Discovery MI 5 scanner at baseline and follow-up. Epicardial adipose tissue (EAT) was quantified from the low-dose non-contrast CT acquired as part of the [$^{18}$F]FDG examination. A pericardial-proxy envelope was constructed from the per-slice 2D convex hull of the cardiac cluster (myocardium, four chambers, proximal aorta, pulmonary artery), dilated 8 mm, and anatomically constrained by the sternum/costal cartilages anteriorly and lung parenchyma laterally. Fat voxels were identified using HU thresholds [−190, −30] (primary) and [−150, −50] (sensitivity analysis). EAT volume was defined as the total fat volume within the envelope minus the cardiac cluster.

#### S3. [$^{18}$F]FDG-PET Quantification

For the head and neck field of view, a single static frame was acquired approximately 30 to 60 minutes post-injection (frame duration: 30 min), reconstructed with motion correction to 1 mm isotropic voxels.

The thoracic field of view was acquired at 60 to 80 minutes post-injection (frame duration: 20 min), reconstructed to 2.0 × 2.0 × 2.8 mm voxels. Two semi-quantitative metrics were computed for all FDG analyses: Standardized Uptake Value (SUV) was calculated as PET tissue concentration (Bq/mL) normalized by body weight and injected dose. Fractional Uptake Rate (FUR) was estimated as the tissue concentration divided by the area under the plasma time-activity curve (AUC) from injection to the scan midpoint, multiplied by 60 to yield units of $min^{-1}$. The plasma input function was constructed by combining an image-derived input function (IDIF) from the descending aorta (0 to 10 min) with manual venous plasma samples (20 to 90 min), integrated using the trapezoidal rule. FUR provides a single-scan proxy for the Patlak net influx rate constant (K*i*) and was selected as the primary FDG metric for the main text because it corrects for inter-individual differences in plasma kinetics. SUV results are reported in Supplementary Table 4.

### S4. Optic Nerve Head (ONH) FDG Analysis

Regions of interest over the left and right optic nerve head were manually delineated on CT images by a trained operator under blinded conditions (randomized session codes). Masks were drawn larger than the anatomical ONH (~1.5 to 2 mm diameter) to ensure coverage given the limited PET spatial resolution (~5.2 mm FWHM at the ONH location). Because the ONH is considerably smaller than the resolution element and mask volumes varied approximately 3.5-fold across delineations, resolution-robust intensity measures were employed: SUVmax (single highest voxel), SUVpeak (mean within a 2 mm radius sphere centered on the maximum), and the mean and 90th percentile of the 150 highest-intensity voxels within each mask ("Top-150"). All masks contained at least 230 voxels, ensuring this metric was unbiased by mask size.

### S5. Periodontal FDG Analysis

Individual teeth were segmented from CT images using TotalSegmentator (v2, teeth task). A peridental soft tissue region of interest was generated for each tooth by dilating the tooth label by 4 mm and subtracting the original tooth volume, creating a shell-shaped ROI. The tongue was segmented separately (TotalSegmentator, head_muscles task) and dilated by 3 mm; this dilated mask was subtracted from all peridental ROIs to minimize contamination from tongue FDG uptake. CT and PET images were co-registered using rigid registration (ANTsPy). Only teeth present in both sessions for a given subject were included, yielding 184 tooth-level observations across 13 subjects (mean 14.2 teeth per subject). Jaw-level metrics were obtained by averaging across all harmonized upper jaw teeth per session. Both weighted mean and 90th percentile (p90) within each ROI were computed.

### S6. Cardiovascular FDG Analysis

The descending thoracic aorta was isolated between vertebral levels T4 and T12 using TotalSegmentator segmentation and connected-component analysis. A vessel wall ROI was constructed by 2D per-slice binary erosion (2 iterations, 4-connected) of the aorta cross-section, yielding an inner-ring shell. Vertebral bodies (dilated 2 mm) and lung parenchyma were excluded. The superior vena cava was eroded by 2 voxels (3D) plus 3 voxels (2D per-slice) to minimize partial-volume effects and served as the blood pool reference region. TBR (tissue-to-background ratio) was then calculated as $SUV_{mean}$ of the vessel wall divided by $SUV_{mean}$ of the blood pool.

**Supplementary Table 1.** Detailed imaging parameters. Additional exploratory outcomes for the different imaging modalities. All p-values from two-sided paired t-tests, uncorrected for multiple comparisons.

| **Outcome** | **Unit** | **n** | **Baseline (mean ± SD)** | **Follow-up (mean ± SD)** | **Δ (95% CI)** | **dz** | **p** |
|---|---|---|---|---|---|---|---|
| ***Heart: Ventricular Volumes and Filling Dynamics*** | | | | | | | |
| LV EDV | mL | 9 | 137.6 ± 40.2 | 142.3 ± 41.4 | 4.65 (−1.52, 10.82) | 0.57 | 0.121 |
| LV ESV | mL | 9 | 49.5 ± 18.4 | 51.4 ± 21.8 | 1.94 (−7.81, 11.68) | 0.15 | 0.659 |
| LV SV | mL | 9 | 88.2 ± 22.7 | 90.9 ± 24.1 | 2.71 (−7.70, 13.12) | 0.20 | 0.564 |
| RV EDV | mL | 9 | 167.4 ± 50.1 | 168.3 ± 47.2 | 0.84 (−7.94, 9.62) | 0.07 | 0.831 |
| RV ESV | mL | 9 | 63.6 ± 21.6 | 61.9 ± 23.8 | −1.77 (−9.79, 6.24) | −0.16 | 0.624 |
| RV SV | mL | 9 | 103.8 ± 29.3 | 106.4 ± 27.9 | 2.61 (−9.39, 14.61) | 0.16 | 0.629 |
| RV EF | % | 9 | 62.3 ± 3.6 | 63.8 ± 6.1 | 1.47 (−3.60, 6.54) | 0.22 | 0.523 |
| Atrial Contribution | % | 9 | −6.9 ± 1.4 | −7.1 ± 1.5 | −0.21 (−1.29, 0.86) | −0.15 | 0.659 |
| Peak Filling Rate | mL/s | 9 | 388.0 ± 101.1 | 470.3 ± 27.4 | 82.26 (−23.03, 87.56) | 0.72 | 0.109 |
| Peak Ejection Rate | mL/s | 9 | 527.8 ± 99.7 | 567.4 ± 38.6 | 39.61 (−78.16, 57.38) | 0.33 | 0.460 |
| ***Heart: 4D Flow LV Blood Flow Routing*** | | | | | | | |
| Direct flow | % | 9 | 43.5 ± 4.2 | 40.0 ± 10.7 | −3.55 (−10.87, 3.76) | −0.37 | 0.295 |
| Retained inflow | % | 9 | 18.9 ± 6.7 | 17.1 ± 7.8 | −1.86 (−5.45, 1.73) | −0.40 | 0.266 |
| Delayed ejection | % | 9 | 17.2 ± 4.9 | 19.3 ± 11.1 | 2.12 (−4.54, 8.78) | 0.24 | 0.483 |
| Residual volume | % | 9 | 20.3 ± 4.2 | 23.6 ± 7.5 | 3.29 (−1.91, 8.48) | 0.49 | 0.183 |
| ***Heart: 4D Flow Kinetic Energy*** | | | | | | | |
| Total average KE | µJ/mL | 9 | 9.2 ± 1.5 | 10.5 ± 2.7 | 1.33 (−1.12, 3.78) | 0.42 | 0.246 |
| Systolic average KE | µJ/mL | 9 | 11.9 ± 2.7 | 13.2 ± 4.7 | 1.38 (−1.88, 4.64) | 0.33 | 0.358 |
| Peak systolic KE | µJ/mL | 9 | 28.6 ± 6.9 | 28.7 ± 10.7 | 0.14 (−8.08, 8.37) | 0.01 | 0.969 |
| Diastolic average KE | µJ/mL | 9 | 7.5 ± 2.2 | 8.7 ± 3.2 | 1.24 (−0.88, 3.36) | 0.45 | 0.213 |
| E-wave average KE | µJ/mL | 9 | 6.5 ± 2.2 | 7.0 ± 2.9 | 0.53 (−1.80, 2.86) | 0.21 | 0.613 |
| E-wave peak KE | µJ/mL | 9 | 15.0 ± 4.5 | 15.7 ± 6.1 | 0.64 (−4.37, 5.65) | 0.12 | 0.776 |
| ***Heart: Vascular FDG and Epicardial Fat*** | | | | | | | |
| Aortic wall TBRmean | — | 13 | 1.14 ± 0.11 | 1.18 ± 0.14 | +0.04 (−0.02, 0.11) | 0.39 | 0.19 |
| Blood pool SUVmean (SVC) | — | 13 | 1.72 ± 0.24 | 1.73 ± 0.32 | +0.01 (−0.15, 0.17) | 0.03 | 0.90 |
| EAT volume (primary) | mL | 13 | 90.2 ± 45.8 | 94.2 ± 45.3 | +3.9 (−3.8, 11.6) | 0.31 | 0.29 |
| EAT volume (sensitivity) | mL | 13 | 62.6 ± 34.8 | 65.7 ± 34.2 | +3.1 (−2.7, 8.8) | 0.32 | 0.27 |

| Outcome | Unit | n | Baseline (mean ± SD) | Follow-up (mean ± SD) | Δ (95% CI) | dz | p |
|---|---|---|---|---|---|---|---|
| ***Retina: RNFL Quadrants (both eyes, n = 26)*** | | | | | | | |
| RNFL Superior | µm | 26 | 110.1 ± 14.9 | 109.3 ± 15.2 | −0.82 (−2.68, 1.05) | −0.16 | 0.377 |
| RNFL Inferior | µm | 26 | 120.2 ± 19.8 | 119.8 ± 17.5 | −0.37 (−2.33, 1.59) | −0.07 | 0.701 |
| RNFL Nasal | µm | 26 | 75.9 ± 11.1 | 76.2 ± 10.5 | 0.29 (−1.55, 2.13) | 0.06 | 0.750 |
| RNFL Temporal | µm | 26 | 64.3 ± 13.9 | 65.1 ± 13.5 | 0.84 (−0.81, 2.49) | 0.18 | 0.305 |
| ***ONH FDG Uptake (FUR, $\times 10^{-3}$ $min^{-1}$)*** | | | | | | | |
| FURmax, Left | $\times 10^{-3}$ | 13 | 16.3 ± 2.5 | 17.4 ± 3.9 | +1.1 (−0.6, 2.8) | 0.40 | 0.18 |
| FURmax, Right | $\times 10^{-3}$ | 13 | 16.7 ± 3.3 | 17.6 ± 3.4 | +0.9 (−1.3, 3.2) | 0.25 | 0.38 |
| FURmax, Bilateral | $\times 10^{-3}$ | 13 | 16.5 ± 2.7 | 17.5 ± 3.5 | +1.0 (−0.6, 2.7) | 0.38 | 0.20 |
| FURpeak, Left | $\times 10^{-3}$ | 13 | 14.5 ± 2.1 | 15.4 ± 3.1 | +0.9 (−0.4, 2.2) | 0.43 | 0.15 |
| FURpeak, Right | $\times 10^{-3}$ | 13 | 14.8 ± 2.7 | 15.6 ± 2.8 | +0.8 (−1.0, 2.5) | 0.27 | 0.35 |
| FURpeak, Bilateral | $\times 10^{-3}$ | 13 | 14.6 ± 2.3 | 15.5 ± 2.8 | +0.8 (−0.4, 2.1) | 0.40 | 0.18 |
| FUR_top150 mean, Left | $\times 10^{-3}$ | 13 | 13.0 ± 1.9 | 14.1 ± 2.4 | +1.1 (+0.1, 2.1) | 0.66 | 0.03 |
| FUR_top150 mean, Right | $\times 10^{-3}$ | 13 | 13.0 ± 2.1 | 14.1 ± 2.4 | +1.0 (−0.2, 2.3) | 0.52 | 0.08 |
| FUR_top150 mean, Bilateral | $\times 10^{-3}$ | 13 | 13.0 ± 2.0 | 14.1 ± 2.3 | +1.1 (+0.0, 2.1) | 0.63 | 0.04 |
| FUR_top150 p90, Left | $\times 10^{-3}$ | 13 | 15.0 ± 2.2 | 16.0 ± 3.1 | +1.0 (−0.4, 2.4) | 0.41 | 0.16 |
| FUR_top150 p90, Right | $\times 10^{-3}$ | 13 | 15.3 ± 2.9 | 16.2 ± 3.0 | +0.9 (−0.9, 2.7) | 0.30 | 0.31 |
| FUR_top150 p90, Bilateral | $\times 10^{-3}$ | 13 | 15.2 ± 2.4 | 16.1 ± 3.0 | +0.9 (−0.4, 2.3) | 0.41 | 0.17 |
| ***L1/L2 Individual Vertebral BMD*** | | | | | | | |
| L1 vBMD | $mg/cm^3$ | 13 | 110.5 ± 25.9 | 113.0 ± 28.2 | +2.46 (−1.98, 6.90) | 0.33 | 0.251 |
| L2 vBMD | $mg/cm^3$ | 13 | 113.3 ± 26.7 | 115.1 ± 27.8 | +1.76 (−2.47, 5.98) | 0.25 | 0.383 |
| ***Muscle Composition*** | | | | | | | |
| Muscle SMD | HU | 13 | 40.7 ± 8.1 | 40.4 ± 8.6 | −0.34 (−2.44, 1.75) | −0.10 | 0.729 |
| Low-density muscle | % | 13 | 30.5 ± 12.3 | 31.4 ± 12.8 | +0.90 (−2.20, 4.00) | 0.18 | 0.539 |
| IMAT | % | 13 | 3.4 ± 2.2 | 3.7 ± 2.8 | +0.33 (−0.21, 0.86) | 0.37 | 0.209 |
| Muscle volume | $cm^3$ | 13 | 206.6 ± 55.7 | 201.5 ± 60.6 | −5.06 (−11.12, 1.00) | −0.50 | 0.094 |
| Muscle CSA | $cm^2$ | 13 | 33.2 ± 9.0 | 32.2 ± 9.7 | −0.97 (−1.98, 0.04) | −0.58 | 0.058 |

*LV = left ventricle; RV = right ventricle; EDV = end-diastolic volume; ESV = end-systolic volume; SV = stroke volume; EF = ejection fraction; KE = kinetic energy; E-wave = early diastolic filling; TBR = tissue-to-background ratio; SVC = superior vena cava; EAT = epicardial adipose tissue; FUR = fractional uptake rate; ONH = optic nerve head; RNFL = retinal nerve fiber layer; vBMD = volumetric bone mineral density; HU = Hounsfield units; IMAT = intermuscular adipose tissue; SMD = skeletal muscle density; CSA = cross-sectional area.*

**Supplementary Table 2.** Exposure-response correlations. Correlations between rapamycin blood concentration (ng/mL; 48h post-dose, week 13) and change in imaging outcomes (Δ = follow-up - baseline).

| Outcome | n | Pearson r | p-value |
|---|---|---|---|
| Δ Cardiac Output | 6 | 0.33 | 0.517 |
| Δ ECV | 6 | 0.01 | 0.984 |
| Δ Peak Filling Rate | 7 | −0.25 | 0.590 |
| Δ Peak Ejection Rate | 7 | 0.43 | 0.335 |
| Δ Perfusion Reserve | 4 | −0.01 | 0.992 |
| Δ Global Longitudinal Strain | 7 | −0.31 | 0.497 |
| Δ A-wave KE, average | 7 | −0.32 | 0.477 |
| Δ A-wave KE, peak | 7 | −0.73 | 0.063 |
| Δ E-wave KE, average | 7 | −0.43 | 0.334 |
| Δ E-wave KE, peak | 7 | −0.58 | 0.173 |
| Δ Direct flow | 7 | −0.13 | 0.786 |
| Δ Retained inflow | 7 | 0.59 | 0.167 |
| Δ Delayed ejection | 7 | 0.14 | 0.770 |
| Δ Pulse Wave Velocity | 7 | –0.30 | 0.508 |
| Δ GCL Thickness | 11 | 0.10 | 0.775 |
| Δ RNFL Thickness | 11 | –0.22 | 0.523 |
| Δ BMD L1–L2 | 11 | 0.29 | 0.39 |
| Δ Muscle CSA | 11 | -0.53 | 0.097 |
| Δ Muscle SMD | 11 | 0.64 | 0.035 |
| Δ Low-density muscle (%) | 11 | -0.59 | 0.058 |
| Δ IMAT (%) | 11 | -0.24 | 0.481 |
| Δ ONH FUR_top150 mean | 11 | 0.26 | 0.43 |
| Δ Periodontal FUR p90 | 11 | 0.06 | 0.865 |
| Δ Aortic wall FURmean | 11 | -0.16 | 0.59 |
| Δ EAT volume | 11 | −0.41 | 0.21 |

**Supplementary Table 3.** Subject completion by modality.

| Subject ID | FDG/CT | Retinal OCT | qCT Bone | Periodontal MRI | Cardiac MRI | PWV |
|---|---|---|---|---|---|---|
| 101 | ✓ | ✓ | ✓ | ✓ | ✓ | ✓ |
| 102 | ✓ | ✓ | ✓ | ✓ | ✓ | ✓ |
| 103 | ✓ | ✓ | ✓ | ✓ | ✓ | ✓ |
| 104 | ✓ | ✓ | ✓ | ✓ | ✓ | ✓ |
| 105 | ✓ | ✓ | ✓ | ✓ | — | — |
| 106 | — | — | — | — | — | — |
| 107 | ✓ | ✓ | ✓ | ✓ | ✓ | ✓ |
| 108 | ✓ | ✓ | ✓ | ✓ | — | — |
| 109 | ✓ | ✓ | ✓ | ✓ | ✓ | ✓ |
| 110 | ✓ | ✓ | ✓ | ✓ | ✓ | ✓ |
| 111 | ✓ | ✓ | ✓ | ✓ | — | — |
| 112 | ✓ | ✓ | ✓ | ✓ | ✓ | ✓ |
| 113 | ✓ | ✓ | ✓ | ✓ | ✓ | ✓ |
| 114 | ✓ | ✓ | ✓ | ✓ | — | — |
| **Total** | **13/14** | **13/14** | **13/14** | **13/14** | **9/14** | **9/14** |

*✓ = completed both baseline and follow-up imaging; — = incomplete or missing data; Subject 106 discontinued treatment early and did not complete follow-up.*

**Supplementary Table 4.** FDG uptake: SUV results. SUV results for all [$^{18}$F]FDG analyses. FUR results are reported in the main text (Table 2) and Supplementary Table 1.

| Metric | Eye | n | Baseline (SD) | Follow-up (SD) | Δ (95% CI) | % Δ | dz | p |
|---|---|---|---|---|---|---|---|---|
| ***ONH FDG: SUV*** | | | | | | | | |
| SUVmax | Left | 13 | 3.00 (0.42) | 3.19 (0.62) | +0.19 (−0.11, +0.49) | +6.3 | 0.39 | 0.19 |
| SUVmax | Right | 13 | 3.08 (0.66) | 3.24 (0.69) | +0.16 (−0.22, +0.54) | +5.3 | 0.26 | 0.37 |
| SUVmax | Bilateral | 13 | 3.04 (0.52) | 3.21 (0.62) | +0.18 (−0.10, +0.45) | +5.8 | 0.39 | 0.18 |
| SUVpeak | Left | 13 | 2.66 (0.37) | 2.82 (0.51) | +0.16 (−0.07, +0.39) | +5.9 | 0.42 | 0.16 |
| SUVpeak | Right | 13 | 2.73 (0.53) | 2.86 (0.55) | +0.13 (−0.16, +0.42) | +4.8 | 0.27 | 0.34 |
| SUVpeak | Bilateral | 13 | 2.69 (0.44) | 2.84 (0.51) | +0.14 (−0.07, +0.36) | +5.4 | 0.41 | 0.17 |
| SUV_top150 mean | Left | 13 | 2.39 (0.34) | 2.58 (0.43) | +0.19 (+0.01, +0.38) | +8.1 | 0.64 | 0.04 |
| SUV_top150 mean | Right | 13 | 2.40 (0.40) | 2.59 (0.43) | +0.18 (−0.01, +0.38) | +7.6 | 0.57 | 0.06 |
| SUV_top150 mean | Bilateral | 13 | 2.39 (0.37) | 2.58 (0.43) | +0.19 (+0.01, +0.36) | +7.9 | 0.66 | 0.04 |
| SUV_top150 p90 | Left | 13 | 2.76 (0.40) | 2.93 (0.53) | +0.17 (−0.09, +0.42) | +6.1 | 0.39 | 0.18 |
| SUV_top150 p90 | Right | 13 | 2.82 (0.55) | 2.98 (0.59) | +0.16 (−0.16, +0.47) | +5.5 | 0.30 | 0.30 |
| SUV_top150 p90 | Bilateral | 13 | 2.79 (0.46) | 2.95 (0.54) | +0.16 (−0.08, +0.40) | +5.8 | 0.41 | 0.16 |

| Metric | n | Baseline (SD) | Follow-up (SD) | Δ (95% CI) | dz | p |
|---|---|---|---|---|---|---|
| ***Periodontal FDG: SUV (Upper Jaw, Jaw-Level)*** | | | | | | |
| SUV mean | 13 | 1.730 (0.278) | 1.797 (0.268) | +0.066 (−0.049, 0.182) | 0.35 | 0.235 |
| SUV p90 | 13 | 2.663 (0.451) | 2.780 (0.419) | +0.117 (−0.062, 0.296) | 0.40 | 0.180 |
| ***Cardiovascular FDG: SUV*** | | | | | | |
| Aortic wall SUVmean | 13 | 1.96 (0.32) | 2.03 (0.36) | +0.07 (−0.09, 0.24) | 0.26 | 0.36 |

*SUV = standardized uptake value; ONH = optic nerve head.. See Supplementary Methods for analysis details.*
